# An experimental evaluation of the relationship between the induced radiofrequency heating near an implanted conductive medical device during MRI, scanner reported B1+rms, and scanner reported average transmit power

**DOI:** 10.1101/2024.03.04.24303732

**Authors:** David H. Gultekin, J. Thomas Vaughan, Devashish Shrivastava

## Abstract

**Background:** Time-varying radiofrequency (RF) fields necessary to perform magnetic resonance imaging (MRI) may induce excessive heating near implanted conductive medical devices during MRI. Both time and space-averaged root mean square of the effective magnetic field (B1+rms) and whole-body average specific absorption rate (SAR) (average RF power per unit body weight) have been proposed as metrics to control the induced heating and avoid unintended thermal injury.

**Purpose:** To evaluate the relationship between the induced RF heating near an implanted conductive medical device, scanner-reported B1+rms, and scanner-reported RF power.

**Methods:** RF heating was measured near the electrodes of deep brain stimulation (DBS) lead placed in a gel phantom using fluoroptic temperature probes in a commercial 3T scanner during MRI. Four transmit and receive RF coil combinations were used, a circularly polarized head transmit and receive coil, a 20-channel head and neck, a 32-channel head, or a 64-channel head and neck receive-only coil with a whole-body transmit coil. RF heating was induced by running a 2D GRE sequence with two RF pulse types (fast and normal) with varying flip angles of 30°, 60°, and 90° and by turning the receive-only coils off and on. The scanner-reported B1+rms and RF power were recorded.

**Results:** Measurements show that the induced temperature change correlated linearly with both the scanner-reported B1+rms and RF power for each coil combination. However, the variation in the induced heating for various RF coil combinations appeared to be much larger for the scanner-reported B1+rms compared to the scanner-reported RF power.

**Conclusion:** Additional studies across other MR scanners are needed to better understand the full extent of the variation in the induced heating near implanted conductive devices as a function of the scanner-reported B1+rms and RF power to develop conservative and reliable patient labeling.

## 1. INTRODUCTION

There are millions of patients implanted with conductive medical devices (1-4). These patients may need to undergo magnetic resonance imaging (MRI) at least once over the lifespan of the device and/or patients for the diagnosis of diseases related or unrelated to the device and/or therapy response monitoring (5-9). However, the interaction between the MR environment and these patients introduces many risks (e.g., displacement, torque, device malfunction, unintended stimulation, etc.) including the possibility of unintended, excessive heating near the implanted devices in a patient, landmark, scanner, and imaging parameters dependent way. The heating may induce thermal damage or deteriorate tissue and/or device function adversely affecting patient safety and device effectiveness (10). The heating is induced because, of the non-uniform, time-varying electromagnetic fields produced in the patient body in and around the device during MRI (11-15).

The International Commission on Non-Ionizing Radiation Protection (ICNIRP) identifies absolute, in vivo local temperature of 4 °C or more or the local temperature change of 5 °C or more in ‘Type-1’ tissues (i.e., all tissues in the upper arm, forearm, and hand, thigh, leg, foot, pinna and the cornea, anterior chamber and iris of the eye, epidermal, dermal, fat, muscle, and bone tissue) and 2 °C or more in ‘Type-2’ tissues (i.e., all tissues in the head, eye, abdomen, back, thorax, and pelvis, excluding those defined as Type-1 tissue) as potentially harmful (16). Per ICNIRP 1998 guidelines “at levels of absorbed electromagnetic energy that cause body temperature rise more than 1–2 °C, a large number of physiological effects have been characterized in studies with cellular and animal systems (17). These effects include alterations in neural and neuromuscular functions; increased blood-brain barrier permeability; ocular impairment (lens opacities and corneal abnormalities); stress-associated changes in the immune system; hematological changes; reproductive changes (e.g., reduced sperm production); teratogenicity; and changes in cell morphology, water and electrolyte content, and membrane functions (18). The International Electrotechnical Commission (IEC) standard “IEC 60601-1:2005/(R)2012 Medical Electrical Equipment – Part 1: General Requirements for Basic Safety and Essential Performance” considers the local absolute temperature of 41 °C as safe (19). The International Organization for Standardization (ISO) standard “ANSI/AAMI/ISO 14708-3:2017 Implants for Surgery – Active Implantable Medical Devices – Part 3: Implantable Neurostimulators” considers the local absolute tissue temperature of 39 °C as safe (20). US Food and Drug Administration (FDA) has accepted the local temperature change near implanted conductive devices of up to 4 °C in thermally non-sensitive, normally perfused muscle-like tissue and up to 2 °C in thermally sensitive, normally perfused muscle-like tissue due to 15 minutes for RF power deposition as safe for imaging patients implanted with the devices for up to an hour in Normal Operating Mode (i.e., the whole-body average SAR of 2 W/kg or less) (21).

Typically, the induced heating is kept below a chosen harmful threshold by keeping the maximum allowable scanner-reported whole-body average specific absorption rate (SAR) (i.e., RF power per unit body weight) and/or time and space averaged root mean square of the effective magnetic field (B1+rms) below a certain value (22-24). Conductive device manufacturers present data acquired experimentally in gel phantoms prepared per voluntary consensus standards and computationally using whole-body human models to regulatory agencies to support patient safety (by keeping expected heating below harmful threshold) for the labeled maximum allowable SAR and/or B1+rms (25,26). The relationship between the scanner-reported RF power and absorbed RF power, and the scanner-reported B1+rms and computational B1+rms are complex. Since, theoretically, the induced heating is proportional to the absorbed power (and not deposited power, scanner reported RF power, or scanner reported B1+rms) for a given implant, implant location, implant medium, and time, to better understand the relationship and/or uncertainty between the induced heating, scanner reported B1+rms, and scanner reported RF power, this novel, preliminary, first-of-a-kind experimental work investigates if i) the induced RF heating near a conductive medical device correlates linearly with the scanner reported B1+rms or scanner reported RF power; and ii) the variation in the induced heating as a function of the scanner reported B1+rms or scanner reported RF power for various RF transmit and receive coil combinations.

## 2. METHODS AND MATERIALS

### 2.1 The Device, Phantom, and RF Heating Measurements

A commercially available and legally marketed deep brain stimulation (DBS) lead was chosen as an example of an implanted conductive device. The lead was placed inside a tissue-mimicking gel phantom prepared per American Society for Testing Materials (ASTM) standard F2182 by using deionized water, 1.32 g/L of sodium chloride (NaCl), and 10 g/L of polyacrylic acid (PAA) (25). The lead was positioned near the sidewalls of a plastic container to maximize the electric field exposure to the device (Figure 1). The connector side of the lead was left open. The induced RF heating was measured by placing calibrated fluoroptic temperature probes (Model: Luxtron m920, Manufacturer: Lumasense Technologies, Fort Collins, CO, USA) near the four electrodes of the lead.

**Figure 1:**
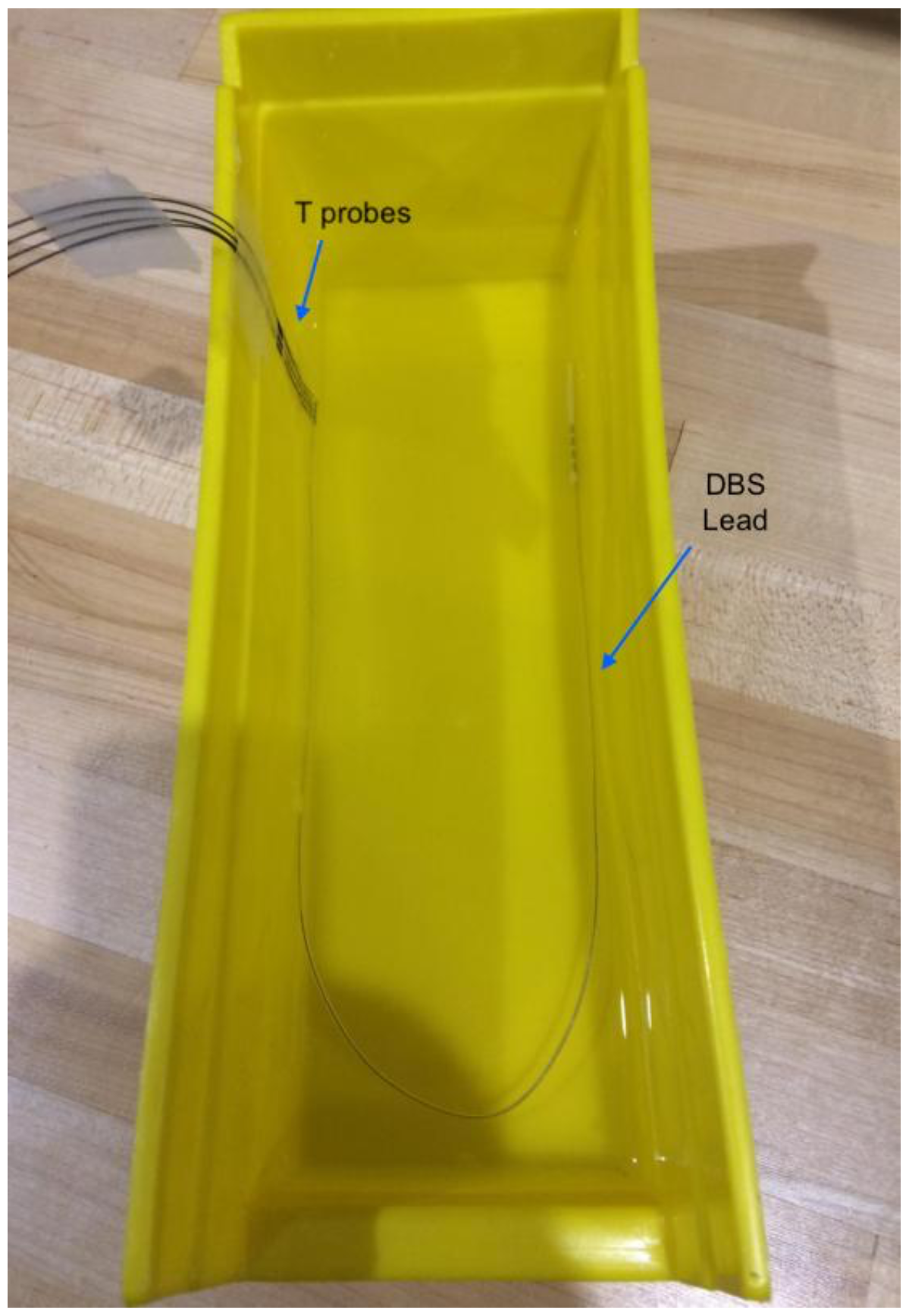
A DBS lead with four electrodes placed inside an ASTM F2182 tissue-mimicking gel. Four fluoroptic temperature (T) probes are placed next to the DBS lead electrodes to measure the induced heating.

### 2.2 Inducing RF Heating

The heating was induced by placing the gel phantom inside a commercial 3 Tesla MRI system and performing imaging with various combinations of RF transmit and receive (Tx/Rx) coils and pulse sequences. Four available RF coil combinations were used, a circularly polarized head transmit and receive (CP Tx/Rx) coil, and a 20-channel head and neck (20 H/N), 32-channel head (32 H), or 64-channel head and neck (64 H/N) receive-only coil with the whole-body transmit coil (BC). RF heating was induced by running a protocol with six sequences, a T_1_-weighted sequence with three flip angles (FA) (30, 60, and 90 degrees), and two RF pulse types (normal, fast) using a two-dimensional (2D) gradient echo sequence (GRE) for each of the RF coil combinations and by turning the receive only coils off and on. The scanner reported B1+rms and power (P) were recorded. No attempt was made to control the scanner reported B1+rms or power (P). Imaging parameters for the 2D GRE sequence were: FA = 30/60/90 degrees, TE = 2.49 ms, TR = 250 ms, TA = 79 s, slice thickness = 4 mm. TR for the fast 2D GRE sequence with 90-degree FA for the 32-channel head receive coil could not be kept at 250 ms and was adjusted by the scanner to 291 ms due to excessive power deposition. It changed the acquisition time for this sequence from 79 s to 91 s. For this sequence, we report the temperature change after 79 s – not after 91 s - to be consistent with other heating results.

## 3. RESULTS

Table 1 presents the induced peak temperature change (dT) near the DBS lead electrodes as a function of the RF coil combinations and pulse sequences. The table shows that the induced heating could vary significantly as a function of RF coil combinations and pulse sequences. The peak temperature change (dT) near the DBS lead electrode is reported at the end of 79 s for all 2D GRE sequences.

**Table 1:**
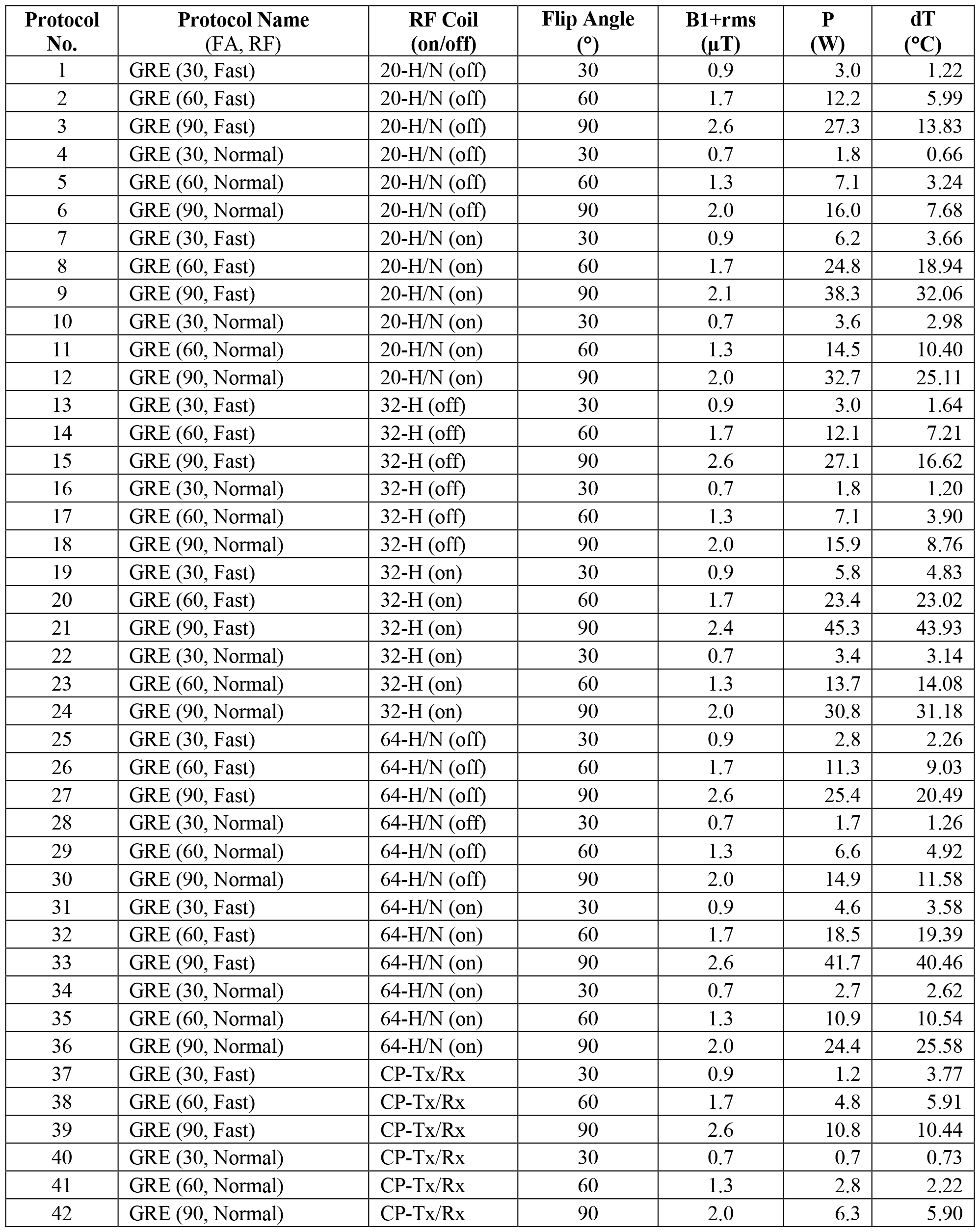
The induced peak temperature change (dT) near the DBS lead electrodes as a function of the RF coil combinations and pulse sequences.

| Protocol No. | Protocol Name (FA, RF) | RF Coil (on/off) | Flip Angle (°) | B1+rms (μT) | P (W) | dT (°C) |
| --- | --- | --- | --- | --- | --- | --- |
| 1 | GRE (30, Fast) | 20-H/N (off) | 30 | 0.9 | 3.0 | 1.22 |
| 2 | GRE (60, Fast) | 20-H/N (off) | 60 | 1.7 | 12.2 | 5.99 |
| 3 | GRE (90, Fast) | 20-H/N (off) | 90 | 2.6 | 27.3 | 13.83 |
| 4 | GRE (30, Normal) | 20-H/N (off) | 30 | 0.7 | 1.8 | 0.66 |
| 5 | GRE (60, Normal) | 20-H/N (off) | 60 | 1.3 | 7.1 | 3.24 |
| 6 | GRE (90, Normal) | 20-H/N (off) | 90 | 2.0 | 16.0 | 7.68 |
| 7 | GRE (30, Fast) | 20-H/N (on) | 30 | 0.9 | 6.2 | 3.66 |
| 8 | GRE (60, Fast) | 20-H/N (on) | 60 | 1.7 | 24.8 | 18.94 |
| 9 | GRE (90, Fast) | 20-H/N (on) | 90 | 2.1 | 38.3 | 32.06 |
| 10 | GRE (30, Normal) | 20-H/N (on) | 30 | 0.7 | 3.6 | 2.98 |
| 11 | GRE (60, Normal) | 20-H/N (on) | 60 | 1.3 | 14.5 | 10.40 |
| 12 | GRE (90, Normal) | 20-H/N (on) | 90 | 2.0 | 32.7 | 25.11 |
| 13 | GRE (30, Fast) | 32-H (off) | 30 | 0.9 | 3.0 | 1.64 |
| 14 | GRE (60, Fast) | 32-H (off) | 60 | 1.7 | 12.1 | 7.21 |
| 15 | GRE (90, Fast) | 32-H (off) | 90 | 2.6 | 27.1 | 16.62 |
| 16 | GRE (30, Normal) | 32-H (off) | 30 | 0.7 | 1.8 | 1.20 |
| 17 | GRE (60, Normal) | 32-H (off) | 60 | 1.3 | 7.1 | 3.90 |
| 18 | GRE (90, Normal) | 32-H (off) | 90 | 2.0 | 15.9 | 8.76 |
| 19 | GRE (30, Fast) | 32-H (on) | 30 | 0.9 | 5.8 | 4.83 |
| 20 | GRE (60, Fast) | 32-H (on) | 60 | 1.7 | 23.4 | 23.02 |
| 21 | GRE (90, Fast) | 32-H (on) | 90 | 2.4 | 45.3 | 43.93 |
| 22 | GRE (30, Normal) | 32-H (on) | 30 | 0.7 | 3.4 | 3.14 |
| 23 | GRE (60, Normal) | 32-H (on) | 60 | 1.3 | 13.7 | 14.08 |
| 24 | GRE (90, Normal) | 32-H (on) | 90 | 2.0 | 30.8 | 31.18 |
| 25 | GRE (30, Fast) | 64-H/N (off) | 30 | 0.9 | 2.8 | 2.26 |
| 26 | GRE (60, Fast) | 64-H/N (off) | 60 | 1.7 | 11.3 | 9.03 |
| 27 | GRE (90, Fast) | 64-H/N (off) | 90 | 2.6 | 25.4 | 20.49 |
| 28 | GRE (30, Normal) | 64-H/N (off) | 30 | 0.7 | 1.7 | 1.26 |
| 29 | GRE (60, Normal) | 64-H/N (off) | 60 | 1.3 | 6.6 | 4.92 |
| 30 | GRE (90, Normal) | 64-H/N (off) | 90 | 2.0 | 14.9 | 11.58 |
| 31 | GRE (30, Fast) | 64-H/N (on) | 30 | 0.9 | 4.6 | 3.58 |
| 32 | GRE (60, Fast) | 64-H/N (on) | 60 | 1.7 | 18.5 | 19.39 |
| 33 | GRE (90, Fast) | 64-H/N (on) | 90 | 2.6 | 41.7 | 40.46 |
| 34 | GRE (30, Normal) | 64-H/N (on) | 30 | 0.7 | 2.7 | 2.62 |
| 35 | GRE (60, Normal) | 64-H/N (on) | 60 | 1.3 | 10.9 | 10.54 |
| 36 | GRE (90, Normal) | 64-H/N (on) | 90 | 2.0 | 24.4 | 25.58 |
| 37 | GRE (30, Fast) | CP-Tx/Rx | 30 | 0.9 | 1.2 | 3.77 |
| 38 | GRE (60, Fast) | CP-Tx/Rx | 60 | 1.7 | 4.8 | 5.91 |
| 39 | GRE (90, Fast) | CP-Tx/Rx | 90 | 2.6 | 10.8 | 10.44 |
| 40 | GRE (30, Normal) | CP-Tx/Rx | 30 | 0.7 | 0.7 | 0.73 |
| 41 | GRE (60, Normal) | CP-Tx/Rx | 60 | 1.3 | 2.8 | 2.22 |
| 42 | GRE (90, Normal) | CP-Tx/Rx | 90 | 2.0 | 6.3 | 5.90 |

Figure 2 shows the distribution of the RF heating as a function of the scanner-reported power for various RF coil combinations and pulse sequences. The induced RF heating appears to vary linearly with the scanner-reported power for various RF coil combinations and all RF coil combinations combined.

**Figure 2:**
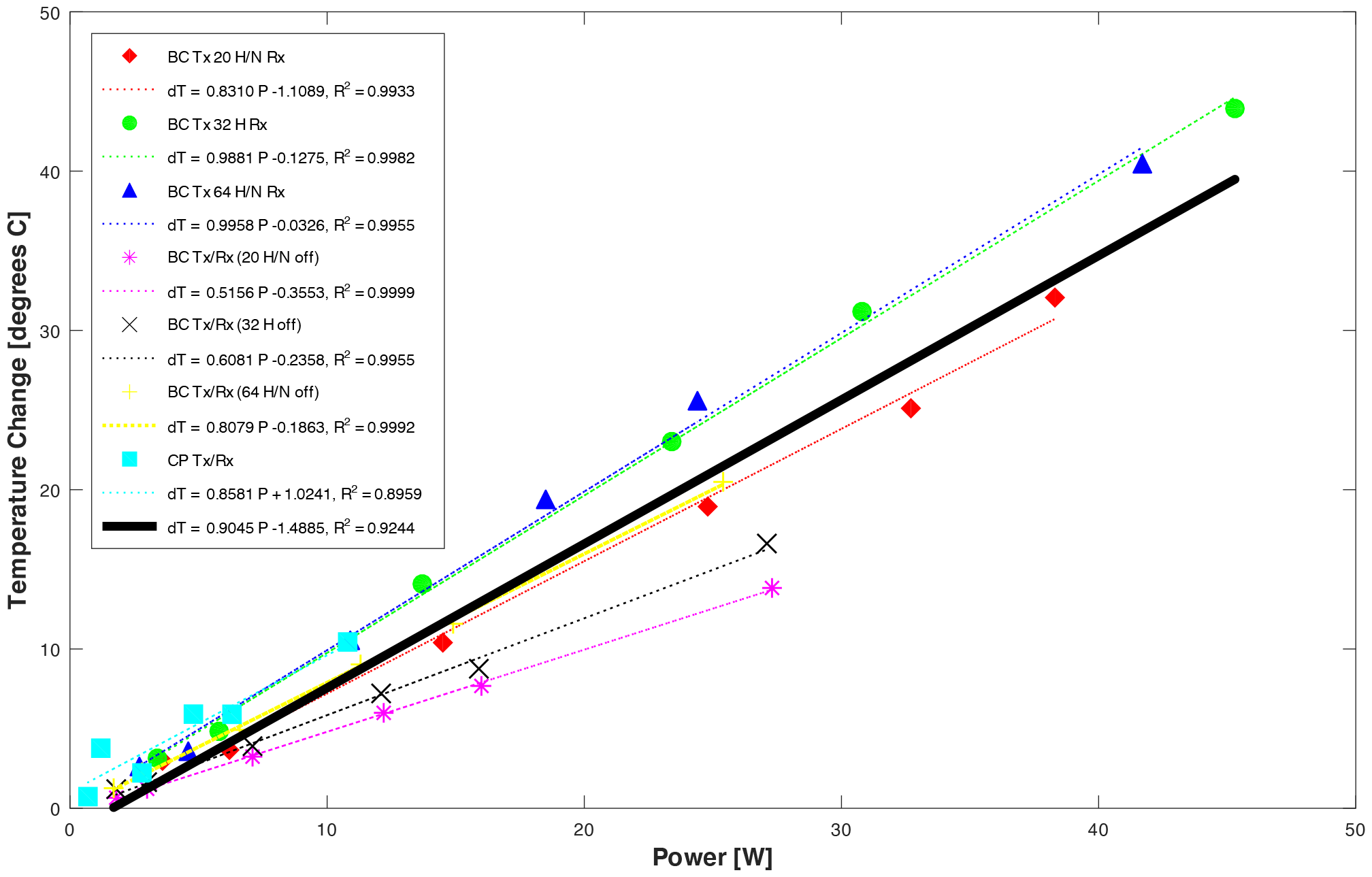
RF heating as a function of the scanner-reported power for all RF coil combinations and pulse sequences.

Figure 3 shows the distribution of the RF heating as a function of the scanner-reported B1+rms for various RF coil combinations and pulse sequences. The induced heating appears to vary linearly with the square of the scanner-reported B1+rms for a given RF coil combination. However, the induced heating appears to vary significantly for all coil combinations combined.

**Figure 3:**
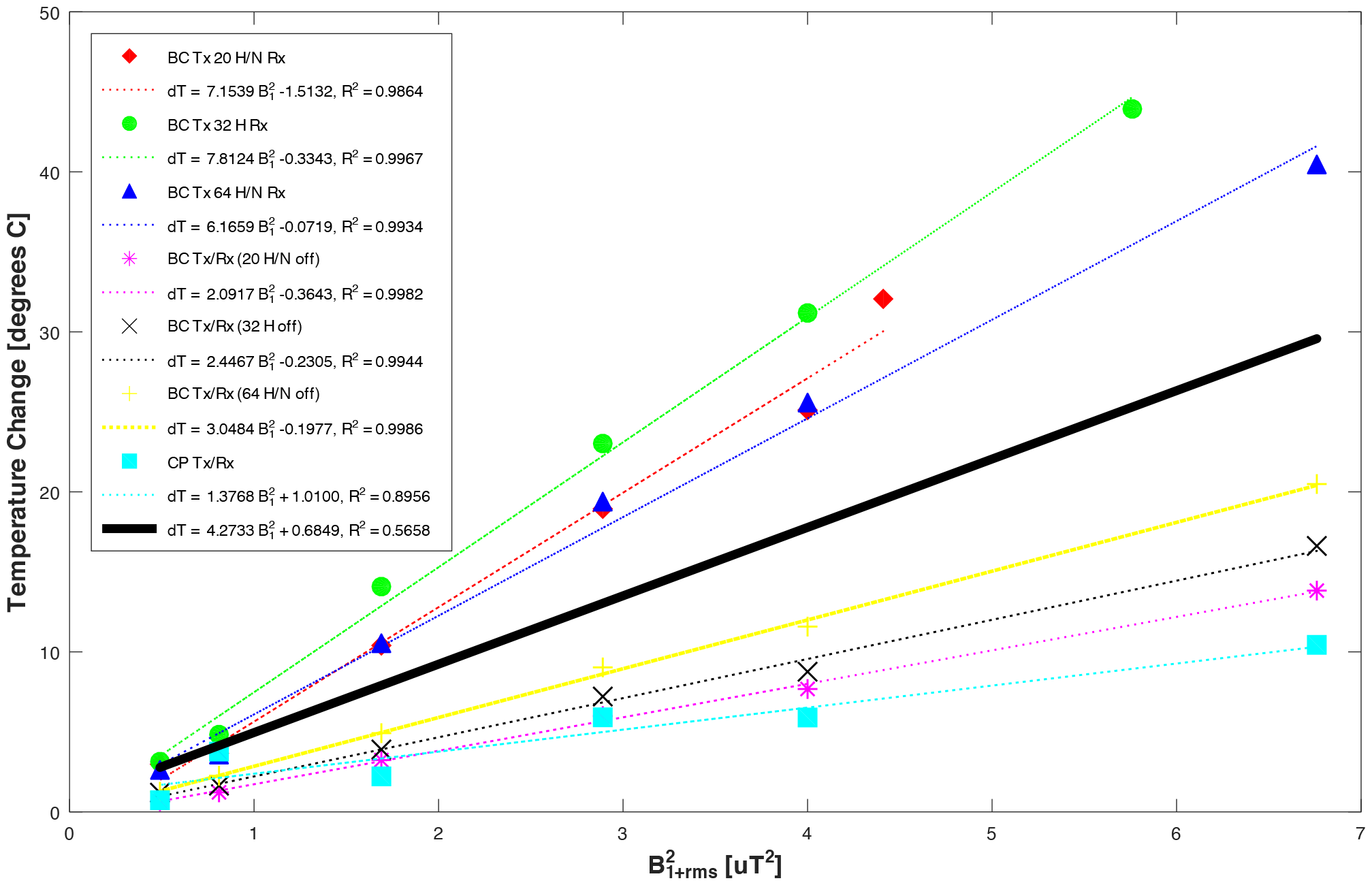
RF heating as a function of the square of the scanner-reported B1+rms for all RF coil combinations and pulse sequences.

Figure 4 presents the distribution of scanner-reported power as a function of the square of the B1+rms for various RF coil combinations and pulse sequences. While, in general, the square of the ratio of the scanner-reported B1+rms was equal to the ratio of the power for varying flip angles and a given RF coil combination, the scanner-reported RF power appeared to vary significantly for a given scanner-reported B1+rms as a function of the RF coil combination.

**Figure 4:**
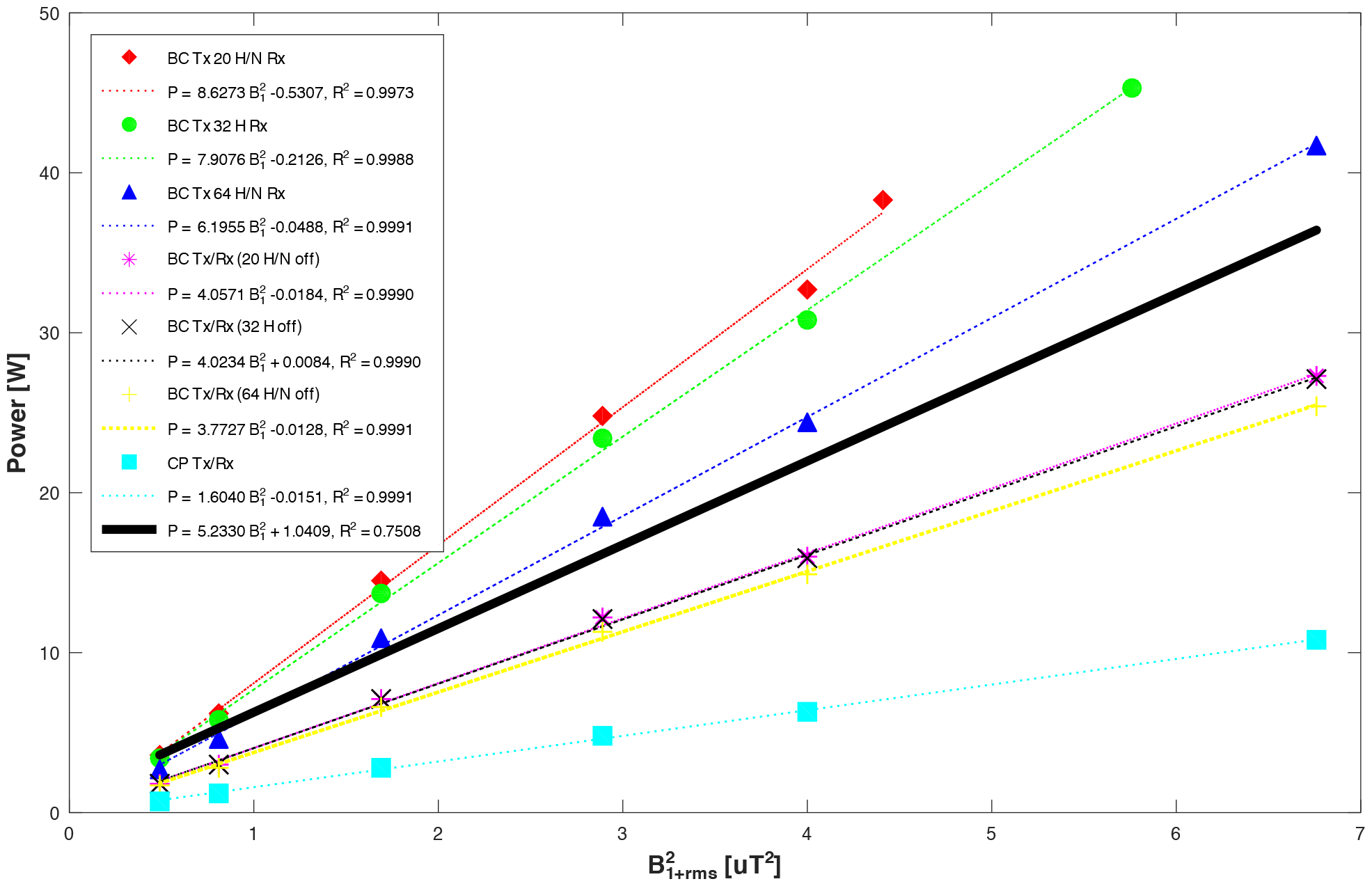
Scanner-reported power as a function of the square of the B1+rms for all RF coil combinations and pulse sequences.

## 4. DISCUSSION

Three main observations were made from the results. First, significant heating may be induced near an implanted conductive medical device during MRI (13,14). This is because of the time-varying, non-uniform electromagnetic fields produced in the human body during MRI-induced currents in the conductive device and heating near the device. The observation suggests the need to accurately determine conditions of safe use to enable imaging patients implanted with conductive devices safely in MRI.

Second, the induced RF heating appears to be linearly correlated with the square of the scanner-reported B1+rms and scanner-reported RF power for a given transmit and receive RF coil combination. This can be explained because the induced RF power density distribution near an implanted device (and thus, the heating) is expected to be linearly related to the RF power delivered from a transmit RF coil and square of the scanner reported B1+rms for a given total RF power loss for a given medium (e.g., gel phantom or human body), implant placement, and medium and implant position in an RF coil combination. The observation suggests that the induced heating may be controlled by regulating the scanner-reported RF power or scanner-reported B1+rms for a given medium, implant placement in the medium, and transmit and receive RF coil combination.

Third, the induced RF heating appears to vary much more for a given scanner-reported B1+rms compared to the scanner-reported RF power for all receive-only and body transmit coil combinations (Figure 2 vs Figure 3). This is because the scanner-reported RF power (and thus the induced heating) appears to vary significantly for a given scanner-reported B1+rms with various receive and body transmit coil, RF coil combinations (Figure 4). The observation suggests that all combinations of transmit and receive coils may need to be studied for a given medium and medium placement within an RF coil combination to identify the transmit and receive RF coil combination that requires depositing maximum power for a given scanner-reported B1+rms to determine the maximum allowable scanner reported B1+rms to keep the induced heating next to implanted conductive devices within the safe threshold. Further, since the induced heating is a function of the local electric field along the device and local electric fields, in turn, are related to local magnetic fields via Maxwell equations, the observation also suggests that it might be important to investigate the relation between the scanner reported B1+rms in a plane to the local magnetic field distribution along the device and heating as a function of the medium and medium placement in an RF coil combination for all RF coil combinations.

Regarding the limitations of the study, the current study elucidates the challenges of developing patient labeling as a function of the scanner-reported B1+rms and/or RF power by keeping the medium, device, device placement inside the medium, and device/medium combination inside an RF coil combination fixed. Since induced RF heating is expected to vary as a function of the medium, device, device placement inside the medium, and device/medium combination inside an RF coil combination, the presented heating results should not be interpreted as a representation of the worst-case heating that could be induced in a gel phantom or in vivo. Finally, the scanner reported B1+rms, real B1+rms in a patient, and computed B1+rms in a whole-body model may vary from one another (27) for different MR manufacturers. Therefore, additional similar experimental studies are needed on MR scanners of various manufacturers to better understand the expected variation in the induced heating near conductive implanted devices for the scanner-reported B1+rms and RF power. Experimental and computational investigations should be conducted together to develop a better and deeper understanding.

## 5. CONCLUSION

The RF-induced heating near an implanted conductive device was shown to correlate linearly with both the scanner-reported B1+rms and RF power for each coil combination. However, the variation in the induced heating for various RF coil combinations appeared to be much larger for the scanner-reported B1+rms compared to the scanner-reported RF power. Additional studies are needed involving various MR scanners to better understand the relationship between the induced heating, B1+rms (scanner-reported B1+rms, real B1+rms in patients, computational B1+rms), and RF power.

## Data Availability

All data produced in the present work are contained in the manuscript

## ACKNOWLEDGEMENTS

This work was supported and performed at the Zuckerman Mind Brain Behavior Institute MRI Platform, a shared resource, and Columbia MR Research Center at Columbia University.

## DISCLAIMER

The subject matter, content, and views presented herein do not represent the views of the Department of Health and Human Services (HHS), the US Food and Drug Administration (FDA), and/or the United States.

